# Using electronic health records to evaluate the adherence to cervical cancer prevention guidelines: a cross-sectional study

**DOI:** 10.1101/2024.03.25.24304195

**Authors:** Kerli Mooses, Aleksandra Šavrova, Maarja Pajusalu, Marek Oja, Sirli Tamm, Markus Haug, Lee Padrik, Made Laanpere, Anneli Uusküla, Raivo Kolde

## Abstract

**Objective:** The fight against cervical cancer requires effective screening together with optimal and on-time treatment along the care continuum. We aimed to examine the impact of cervical cancer screening and treatment guidelines on screening, and follow-up adherence to guidelines.

**Methods:** Data from electronic health records and healthcare provision claims for 50 702 women was used. The annual rates of PAP tests, HPV tests and colposcopies during two guideline periods (2nd version 2012–2014 vs 3rd version 2016–2019) were compared. To assess the adherence to guidelines, the subjects were classified as adherent, over- or undertested based on the timing of the appropriate follow-up test.

**Results:** The number of PAP tests decreased and HPV tests increased during the 3rd guideline period (p < 0.01). During the 3rd guideline period, among 21–29-year-old women, the adherence to guidelines ranged from 38.7% (44.4…50.1) for ASC-US to 73.4% (62.6…84.3) for HSIL, and among 30–59-year-old from 49.0% (45.9…52.2) for ASC-US to 65.7% (58.8…72.7) for ASC-H. The highest rate of undertested women was for ASC-US (21–29y: 25.7%; 30–59y: 21.9%). The rates of over-tested women remained below 12% for all cervical pathologies observed. There were 55.2% (95% CI 49.7…60.8) of 21–24-year-old and 57.1% (95% CI 53.6…60.6) of 25–29-year-old women who received an HPV test not adherent to the guidelines.

**Conclusions:** Our findings highlighted some shortcomings in the adherence to guidelines, especially among women under 30. The insights gained from this study helps to improve the quality of care and thus, reduce cervical cancer incidence and mortality.

## INTRODUCTION

Cervical cancer, one of the most common malignant neoplasms among women [1], is preventable with the timely discovery of pre-cancerous lesions and appropriate treatment [2]. Moreover, World Health Organisation has set a target to eliminate cervical cancer as a public health problem [2]. To fulfil this goal, countries have developed their own national guidelines for screening and management of cervical abnormalities, while others adhere to only international ones [8,9]. These guidelines have changed over time together with the evolution of scientific knowledge [3] and try to offer the best protection against cervical cancer while balancing the benefits and harms of the testing and ensuring the effective use of healthcare resources. Therefore, it is crucial that these guidelines are followed and effectively implemented in the practice. Despite this, both underscreening and over-screening has been reported [4–8]. For example, underscreening has remained a challenge in several countries as the adherence to screening guidelines in developed countries is on average 63% [4], ranging from 37% to 82% in Europe [5]. At the same time, a considerable amount of women are tested more often than recommended [6,7,9]. Too frequent testing is not only a financial burden to the healthcare system but can lead to overtreatment and have a negative impact on the reproductive health [10]. While several studies have found difficulties in adopting new guidelines in the US and thus, causing the increase in over-testing [6,7,9], there is a lack of similar studies in Europe.

To assure screening benefits, it is crucial to involve and maintain women from screening to treatment and to follow-up along a care continuum. It has been estimated that 12% of invasive cervical cancer cases are attributable to poor follow-up after abnormal PAP test [11]. To date, the data about treatment and follow-up patterns is limited and the need for additional studies has been stressed [12,13]. There are some studies describing the follow-up of abnormal screening results, however, in most cases, the focus is on the timing of the follow-up test [12,14–16], while less attention has been paid to the additional tests [13]. More detailed information about the treatment pathways help to provide better treatment and care for women and reduce the incidence of cervical cancer.

Previously, it has been highlighted that there is a considerable amount of research that fails to improve practice as it does not address questions that are important to practitioners, it is not applicable to real-life, patient-centred or available to policy makers [17–20]. To increase the practicability of current study and to ensure that the results are disseminated and implemented into the practice, it has been conducted in close co-operation with Estonian Gynaecologists Society and several practitioners. Moreover, to get better overview of the real situation we incorporated diverse types of real-world data. Thus, in the current study, we utilized nationwide and population-based electronic health record (EHR) and healthcare provision claims data with the aim to investigate the influence of cervical cancer screening guidelines on testing practices, and the adherence to guidelines in the case of follow-up tests.

## MATERIAL AND METHODS

### Setting

In Estonia, the evidence-based guideline for diagnosing, monitoring, and treatment of precancerous cervical, vaginal, and vulvar conditions is developed and disseminated by the Estonian Gynaecologists Society. In the current cross-sectional study, we focus on the 2^nd^ (2007–2014) and 3^rd^ (2014–2021) versions of the guidelines. During the 2^nd^ version the primary screening test was the PAP test with a one-year interval between the first two tests, which in case of no abnormalities, was followed by a two-year testing interval. The 3^rd^ version of the guideline recommended a three-year testing interval with a PAP test or a five-year interval when HPV or co-testing was applied.

### Data

In this cross-sectional study we combined data from two health databases with national coverage – EHR and healthcare service provision claims from 01.01.2012 to 31.12.2019. The databases were linked using a unique personal code given to all persons living in Estonia. The data was transferred to Observational Medical Outcomes Partnership (OMOP) common data model (CDM) version 5.3 [22] described in more detail by Oja et al [23]. The study sample consisted of randomly selected 10% of the Estonian female population aged 16–65 years (N = 50 702) and followed the age distribution of the whole female population [23]. The study was approved by the Research Ethics Committee of the University of Tartu (300/T-23) and the Estonian Committee on Bioethics and Human Research (1.1-12/653) and the requirement for informed consent was waived.

The episodes of cervical cytology together with diagnoses, HPV tests and colposcopies were identified from EHR and claims databases. When one PAP test had multiple diagnoses (eg NILM and LSIL), the most severe pathology was used. Cervical cytology results were categorised according to the Bethesda system criteria:[24] negative for intraepithelial lesion or malignancy (NILM); atypical squamous cells of undetermined significance (ASC-US); low-grade squamous intraepithelial lesions (LSIL); atypical squamous cells cannot exclude high-grade squamous intraepithelial lesions (ASC-H); high-grade squamous intraepithelial lesions (HSIL). The corresponding Standard Nomenclature of Medicine (SNOMED) codes are presented in Table S1.

### Assessing the effect of guidelines on the testing practices

The data about all PAP tests, HPV tests and colposcopies during 2012–2014 (2^nd^ version) were compared with data from 2016–2019 (3^rd^ version). The year 2015 was considered a transition period and was left out from the comparison. In this analysis, we present the annual testing rates for the target group of the guidelines (21–65-year-old women) and the younger age group (16–20-year-old) who should not receive any testing.

### Assessing the adherence to cervical cancer prevention and care continuum

#### Follow-up of PAP tests

We described the follow-up during the 3^rd^ guideline version (2015–2019) when the PAP result was NILM, ASC-US, LSIL, ASC-H, or HSIL. For each PAP test result under observation a separate cohort was created with similar inclusion criteria: 1) the first corresponding PAP test result was recorded after 01.01.2015 (index event), 2) there was no occurrence of more serious cervical pathology 30 days before and after the index date, 3) no previous diagnosis of the same cervical pathology within three years before the index event was present (not applicable to NILM), 4) a follow-up period after the index date was at least 15 months for cervical pathologies and 3.5 years for NILM. Women with a history of HIV, cervical cancer or hysterectomy were excluded from the analysis. When describing the follow-up of NILM we also excluded women whose PAP test was accompanied by an HPV test, as in the case of co-testing, the recommended testing interval was longer (five years). In this analysis we focused on two age groups: 21–29 and 30–59-year-olds.

In Table 1 we present the follow-up tests after NILM and cervical pathologies according to the guidelines together with the adherence and undertested criteria deployed in the analysis. The adherence to guidelines was assessed in terms of timing and appropriate follow-up test. When the appropriate follow-up test was performed in a time interval specified in the guidelines and no additional test was present, the woman was considered as adherent. To provide some flexibility and account for possible shifts caused by appointment scheduling, we allowed a time lag when classifying the test as adherent. For example, according to the guidelines in the case of NILM the follow-up PAP test should be done after three years, however, when the next PAP test was done two years and seven months or three years and four months after the index date, it was considered adherent. Those women who had a follow-up test according to the guidelines but in addition had some PAP or HPV test(s) between the index test and an adherent follow-up test, were classified as over-tested, while women who did not receive any test after the PAP test, were classified as undertested. Women who did not belong to adherent, over- or undertested group were left out from current analysis (n = 2997, 32.5%).

**Table 1.** The follow-up tests of NILM and cervical abnormalities according to the guidelines and the adherence definition in the analysis.

|  | Age | Guidelines | Adherent in the analysis* | Undertested in the analysis |
| --- | --- | --- | --- | --- |
| NILM | 21–59 | New PAP test after 3 years | New PAP test between 2.5 to 3.5 years | No test within 3.5 years |
| ASC-US | 21–59 | New PAP test after 12 months<br>OR<br>Immediate HPV test | New PAP test between 9–15 months<br>OR<br>HPV test in 3 months | No test within 15 months |
| LSIL | 21–24 | New PAP test after 12 months | New PAP test between 9–15 months | No test within 15 months |
|  | 25–59 | New PAP test after 12 months<br>OR<br>Immediate colposcopy | Co-test between 9–15 months<br>OR<br>Colposcopy in 3 months |  |
| ASC-H | 21–59 | Colposcopy | Colposcopy in 3 months | No test within 12 months |
| HSIL | 21–59 | Colposcopy | Colposcopy in 3 months | No test within 12 months |

\* no additional test between index and follow-up test

#### HPV testing among women aged 21–29 years

The HPV test was not recommended as a screening test in this age group. However, in the case of cervical pathology ASC-US among 21–24-year-old women and ASC-US or LSIL among 25–29-year-old women, HPV test was allowed. Thus, these age groups were analysed separately. The index date was the first HPV test between 01.01.2016–31.12.2019. Next, the preceding PAP test together with its result was identified. The period for the preceding PAP test was set 12 months before or 30 days after the index date.

### Data analysis

To describe the overall sample, we calculated the annual rates of PAP tests, HPV tests and colposcopies per 100 000 women together with 95% confidence intervals (95% CI) using the midyear population counts for the denominator. Repeated testing within 30 days was considered duplicates and only the first record was included in the analysis. The threshold of 30 days was chosen to account for different testing and laboratory analysis pathways. The differences in the number of tests between the two guideline periods were described using absolute and relative change of rates. The p-values were calculated using a z-test of proportions, and the statistical significance level was set at p < 0.05. When describing the adherence, under- and over-testing during the 3^rd^ guideline period, ratios together with 95% CI are presented. The analysis was performed using R v4.1.2.

## RESULTS

### The study cohort

During the study period, 72.6% of 21–65-year-old and 23.7% of 16–20-year-old women had received at least one PAP test (Table 2).

**Table 2.** PAP tests, HPV tests, and colposcopies in 2012–2019 for 21–65-year-old (n = 47 570) and 16–20-year-old (n = 7 404) women, in Estonia. The number of tests, women who received at least one test and their proportion.

| Age: | Procedures (n) |  | Women received at least one procedure (n (%)) |  |
| --- | --- | --- | --- | --- |
|  | 16–20y | 21–65y | 16–20y | 21–65y |
| PAP test | 2 567 | 103 213 | 1 755 (23.7%) | 34 530 (72.6%) |
| HPV test | 236 | 10 177 | 206 (2.8%) | 7 060 (14.8%) |
| Colposcopy | 178 | 6 830 | 135 (1.8%) | 3 972 (8.3%) |

### The effect of guidelines on the testing practices

During the 3^rd^ guideline period (2016–2019) there was a statistically significant decline in the number of PAP tests and an increase in HPV tests in both age groups compared with the 2^nd^ guideline version (*P* < 0.001) (Table S2). The decline in colposcopies during the 3^rd^ period was present only among 16–20 years-old women (*P* < 0.01) (Table S2). The annual PAP testing, HPV testing and colposcopy rates per 100 000 women is presented in Figure 1.

**Figure 1.**
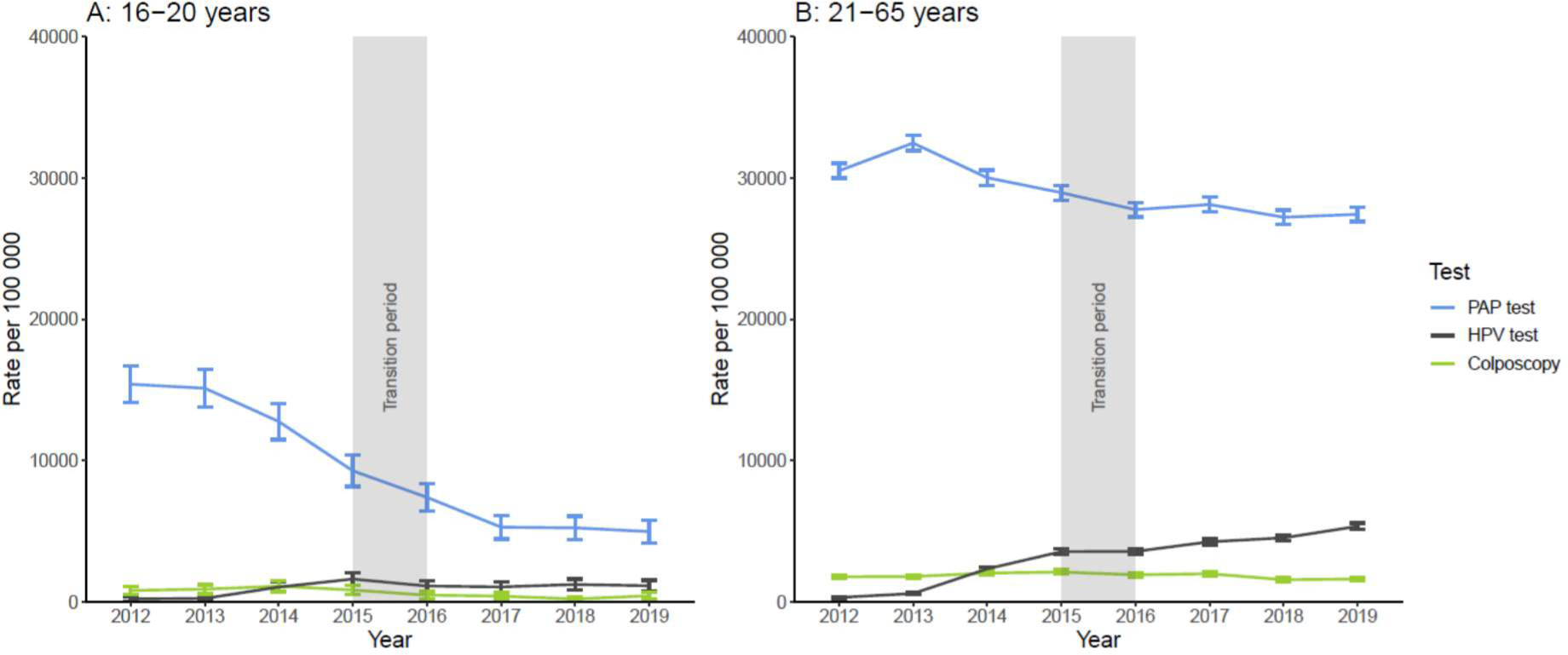
The rate of women per 100 000 who received PAP tests, HPV tests or colposcopies during 2^nd^ and 3^rd^ guideline versions (2012–2014 vs 2016–2019) in the age groups 16–20 years (A) and 21–65 years (B).

### Adherence to cervical cancer prevention and care continuum in 2015–2019

The adherence to guidelines after NILM was 22.7% (95% CI 20.4 - 24.8) among 21–29-year-old and 20.5% (95% CI 19.4 - 21.5) among 30–59-year-old women (Table 3).

**Table 3.**
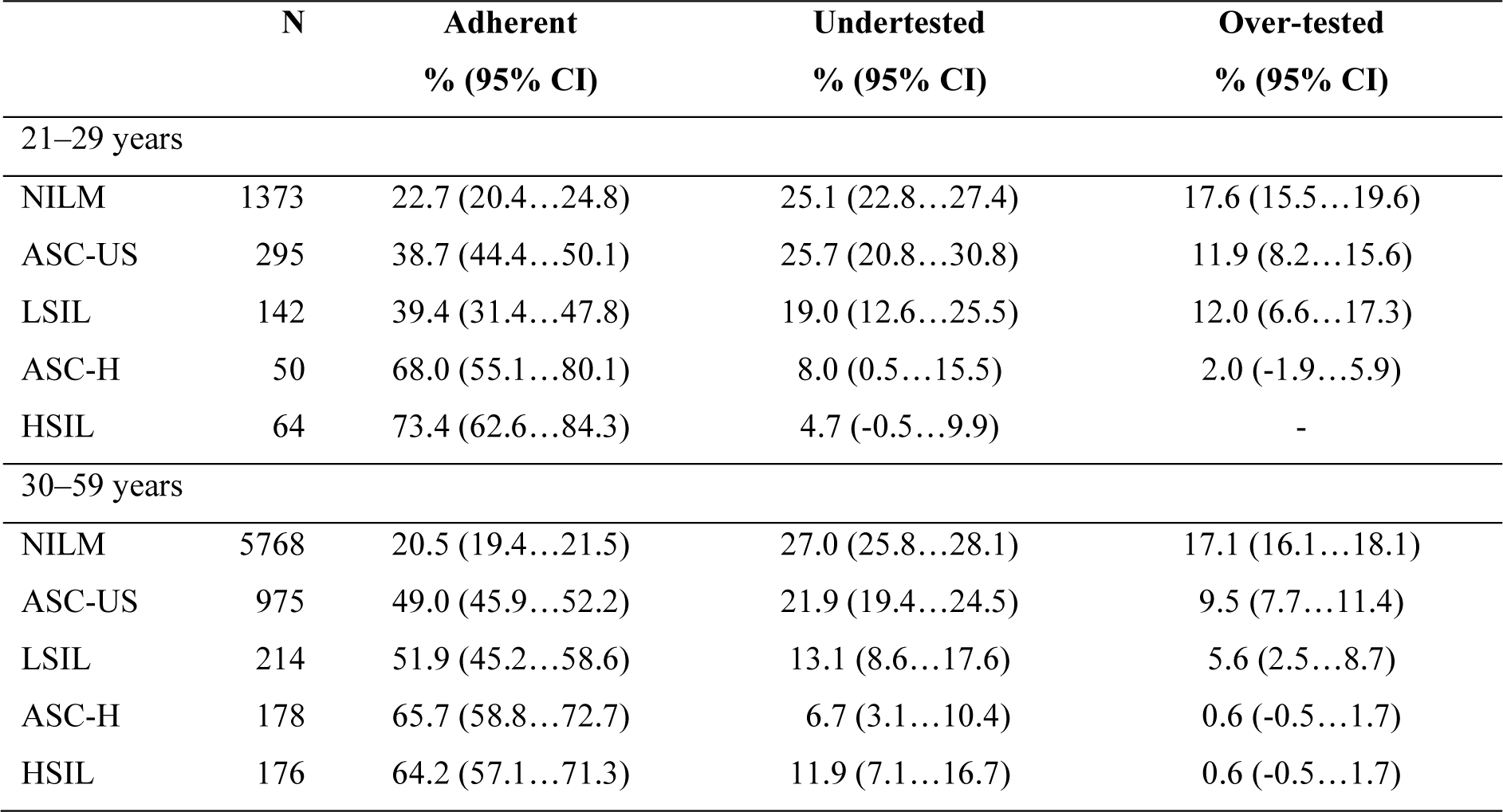
The proportion of adherent, under- and over-tested women based on the PAP test outcome among age groups 21–29 and 30–59 years.

Among 21–29-year-old women, 68.0% (95% CI 55.1 - 80.1) of women with ASC-H and 73.4% (95% CI 62.6 - 84.3) with HSIL were adherent with the guidelines (Table 3). When looking at a one-year period, the proportion of 30–59-year-old women receiving colposcopy was 80.0% (95% CI 68.9 - 91.1) for ASC-H and 87.5% (95% CI 79.4 - 95.6) for HSIL (Figure 2). The proportion of women with no follow-up activity after ASC-H and HSIL was below 10% (Table 3). In the case of milder pathologies (ASC-US, LSIL) less than half of 21–29-year-old women were adherent with the guidelines, while 25.7% (95% CI 20.8 - 30.8) of women with ASC-US and 19.0% (95% CI 12.6 - 25.5) of women with LSIL did not receive any tests within 15 months after the first corresponding pathology.

**Figure 2.**
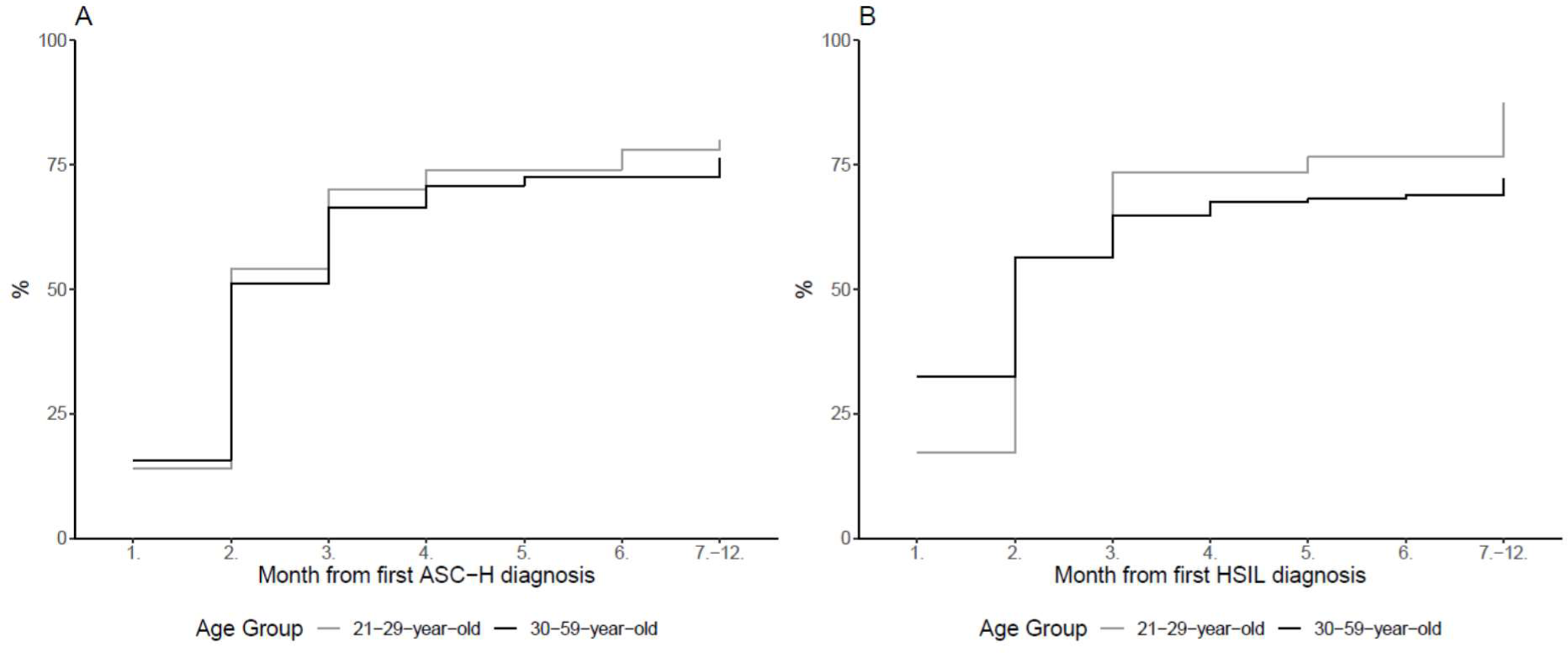
The time of colposcopy after ASC-H (A) and HSIL (B) by age group.

As for 30–59-year-old women, in the case of cervical pathology the adherence to guidelines ranged from 49.0% (95% CI 45.9 - 52.2) for ASC-US to 65.7% (95% CI 58.8 - 72.7) for ASC-H (Table 3). The highest rate of undertested women with a cervical pathology was observed for ASC-US (21.9%, 95% CI 19.4 - 24.5) and lowest for ASC-H (6.7%, 95% CI 3.1 - 10.4). When looking at a one-year period, 76.4% (95% CI 70.2 - 82.6) of women with ASC-H and 72.2% (95% CI 65.5 - 78.8) with HSIL received a colposcopy (Figure 2). As for the HPV testing among women aged 21–29 years there were 55.2% (95% CI 49.7 - 60.8) of 21–24-year-old and 57.1% (95% CI 53.6 - 60.6) 25–29-year-old women who received an HPV test without a preceding PAP test or the result of the preceding PAP test did not require additional HPV test (Figure 3). The proportion of women whose HPV test was performed according to the guidelines was 18.0% (95 CI 13.7 - 22.3) among 21–24-year-old and 26.6% (95% CI 23.5 - 29.7) among 25–29-year-old women.

**Figure 3.**
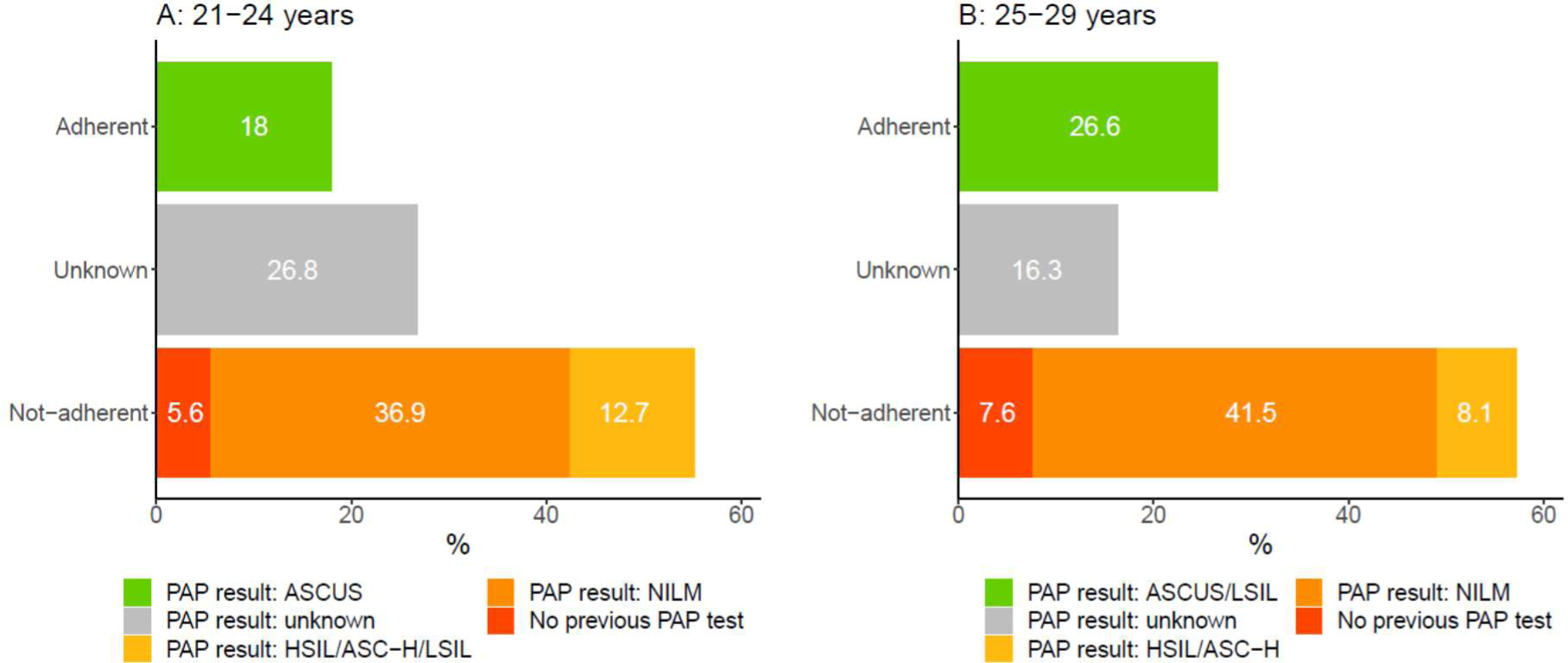
The distribution of 21–24 (A) and 25–29 (B) year-old women who had their first HPV test between 2016–2019 based on the preceding PAP test.

## DISCUSSION

The study extends the current literature on the adherence to cervical cancer screening guidelines. Our findings indicate that the guidelines have an influence on the practice, and that overall, the new guideline has been adopted by the practitioners. Still, there are some shortcomings that need to be addressed. For example, the adherence rates of follow-up ranged from 40–73% depending on the cervical pathology and age, while better follow-up care was provided for women with high-grade pathology. A considerable proportion of women were undertested and thus, did not receive an optimal protection against cervical cancer, while overtesting was more prevalent among young women under 30. Our results give a good insight into the quality of care and highlight aspects that need to be addressed to reduce the incidence and mortality of cervical cancer.

After implementing the 3^rd^ version of the cervical cancer screening guidelines, the number of PAP tests reduced and HPV tests increased. These trends in PAP tests in both age groups and HPV tests among 21–65-year-old women are consistent with previous studies [8,24,25]. Although some studies have suggested that increasing the testing interval can be a barrier to implementing the guidelines [6,9], this seems not to be the case in our study. It could be hypothesised that the relatively successful adoption of the new guideline can, at least partly, be attributable to the fact that the national gynaecologists society led the whole guideline development and dissemination process.

However, our study documented considerable over-testing. First, there was a significant increase in HPV testing among 16–20-year-old women during the 3^rd^ guideline version compared to the previous guideline. As HPV test is not recommended in this age group, this trend can be classified as over-testing. This increase contradicts earlier studies conducted in the US where the number of HPV tests remained steady [25] or had a slight decline [25]. Secondly, we observed a substantial over-testing among 21–29-year-old women where more than half of HPV tests did not have a preceding PAP test result as recommended by the guidelines. Thirdly, 12% of young women with ASC-US and LSIL and a fifth with NILM received additional tests before the follow-up test recommended by the guidelines. Therefore, the over-testing tends to be more prevalent among women under 30. Such surplus tests are not only an inefficient use of healthcare resources, but they can cause stress and anxiety for women [27] and negatively impact the reproductive health [10]. Moreover, as the low-grade cytologic findings of young women tend to experience regression without any intervention [10,28], caution is recommended in treating them [29]. It could be hypothesised that the increase in HPV testing among young women is due to the guideline change as HPV test became the primary screening test in the 3rd guideline and the recommended age range was just unnoticed by the practitioners. Our findings suggests that more effort should be made to raise the awareness of clinicians about the recommended testing and treatment pathways among young women.

In accordance with the previous findings [12], our study indicates that women with high-grade cervical pathology had better follow-up care than low-grade pathology. This is reflected both in the adherence rates as well as in the proportion of undertested women. The adherence rates for high-grade cervical pathologies (ASC-H and HSIL) ranged in our study from 64% among 30–59-year-old with HSIL to 73% among 21–29-year-old with HSIL, which were lower than previously reported in Canada [16] and higher than in the USA [29]. Whereas the adherence rates for low-grade cervical pathologies in our study remained around 39% to 52%. However, it should be highlighted, that despite having a better follow-up care in the case of high-grade cervical pathology, almost a fifth of women with ASC-H and HSIL did not receive a colposcopy within one year. When looking at the proportion of undertested women, the rates of women without any follow-up test were a higher for women with low-grade cervical pathology. Somewhat surprisingly, the lowest proportion of adherent women was in the case of NILM, where only a fifth of women received their next PAP test as recommended in the guidelines, while a quarter of women did not receive any additional testing within three and half years. The proportion of adherent women with NILM in the current study is significantly lower, and the proportion of undertested women higher compared to previously reported [31,32]. Although irregular testing is better than no testing [33,34], the best protection against cervical cancer is provided with regular testing and appropriate follow-up of cervical pathologies [35,36]. Previously it has been shown that the main reason for under-screening is the lack of knowledge of the guidelines among women [37]. Our results highlight the need to improve the overall knowledge about cervical cancer and also an overall health literacy of women. It has been pointed out that difficulty in understanding health information, lower knowledge and worry about cervical cancer contribute to nonadherence to cervical cancer screening [37,38]. In contrast, according to a meta-analysis, better health literacy is associated with higher cervical cancer screening participation [40]. In addition, there are several organizational factors that can support the participation in screening, such as receiving personal reminders via different channels [41,42], or making the participation as convenient as possible.

It has been shown that long waiting lists, inappropriate appointment times, and distance from the clinic are some reasons for nonadherence [43]. Therefore, a multidisciplinary approach is needed where both the individual as well as healthcare system are addressed.

When interpreting our study results some limitations should be considered. First, there are some completeness issues as 45% of the PAP test had no result reported in the EHR which hinders the long-term analysis of testing patterns. Therefore, our study focused only on the first follow-up test rather than the whole treatment pathway. Second, the short timespan under observation set some restrictions on evaluating the over- and undertesting, which could give a better understanding of implementing the guidelines. Third, the study is also limited by the lack of information on socio-demographic variables. At the same time, the strength of current study is the close co-operation with Estonian Gynaecologists Society and clinical practitioners which helped to identify several knowledge gaps in the implementation of cervical cancer prevention guidelines. We addressed these gaps using real-world data by combining EHR and claims database which store healthcare data for almost all Estonians, both insured and uninsured [23]. The results of the study will be used to improve the implementation of the guidelines, plan targeted interventions and nudge the policy makers and other stakeholders to support the effective implementation of the guidelines to provide optimal treatment and care for the women which in long-term should also result in the reduced incidence and mortality of cervical cancer.

## CONCLUSION

The study gave an important insight of the actual clinical practice and helped to identify several shortcomings in the implementation of cervical cancer prevention guidelines that need to be addressed. The practical input of the study will be used to provide better optimal treatment and better care for the women on national level. For some pathologies and age groups the adherence, overtesting or undertesting patterns seem to be similar internationally, indicating universal trends to be present. However, there is a lack of studies monitoring the guideline adherence for follow-up testing. Thus, more research is needed to reveal both unnecessary and insufficient testing practices and take action against them.

## Data Availability

There are legal restrictions on sharing de-identified data. According to legislative regulation and data protection law in Estonia, the authors cannot publicly release the data received from the health data registries in Estonia.

## Abbreviations

AIS: adenocarcinoma in situ of the cervix
ASC-H: atypical squamous cells cannot exclude HSIL
ASC-US: atypical squamous cells of undetermined significance
EHR: electronic health record
HSIL: high-grade squamous intraepithelial lesions
LSIL: atypical squamous cells of undetermined significance
NILM: negative for intraepithelial lesion or malignancy
OMOP: Observational Medical Outcomes Partnership

## DECLARATIONS

### Ethics approval and consent to participate

The study was approved by the Research Ethics Committee of the University of Tartu (300/T-23) and the Estonian Committee on Bioethics and Human Research (1.1-12/653) and the requirement for informed consent was waived.

### Consent for publication

Not applicable.

### Authors’ contribution

All authors participated in the conceptualisation, planning, revision and editing process and have approved the submitted version. In addition, data, the methodology was developed by KM, MO, ST, AU, RK. The data cleaning, data analysis and visualisations were carried out by MO, ST, MH, KM, MP. The initial draft was written by KM.

## Acknowledgements

Not applicable.

